# Binding and Neutralizing Antibody Responses to SARS-CoV-2 in Infants and Young Children Exceed Those in Adults

**DOI:** 10.1101/2021.12.20.21268034

**Authors:** Ruth A. Karron, Maria Garcia Quesada, Elizabeth A. Schappell, Stephen D. Schmidt, Maria Deloria Knoll, Marissa K. Hetrich, Vic Veguilla, Nicole Doria-Rose, Fatimah S. Dawood, SEARCh Study Team

**Author notes:** **Disclaimers:** The findings and conclusions in this report are those of the authors and do not necessarily represent the views of the U.S. Centers for Disease Control and Prevention. Walker Black^3^, Christine Council-DiBitetto^1^, Tina Ghasri^1^, Amanda Gormley^1^, Milena Gatto^1^, Maria Jordan^1^, Karen Loehr^1^, Jason Morsell^1^, Jennifer Oliva^1^, Jocelyn San Mateo^1^, Khadija Smith^1^, Kimberli Wanionek^1^, Cathleen Weadon^1^, Suzanne Woods^1^.

## Abstract

SARS-CoV-2 infections are frequently milder in children than adults, suggesting that immune responses may vary with age. However, information is limited regarding SARS-CoV-2 immune responses in young children. We compared Receptor Binding Domain binding antibody (RBDAb) and SARS-CoV-2 neutralizing antibody (neutAb) in children aged 0-4 years, 5-17 years, and in adults aged 18-62 years in a SARS-CoV-2 household study. Among 55 participants seropositive at enrollment, children aged 0-4 years had >10-fold higher RBDAb titers than adults (373 vs.35, *P*<0.0001), and the highest RBDAb titers in 11/12 households with seropositive children and adults. Children aged 0-4 years had 2-fold higher neutAb than adults, resulting in higher binding to neutralizing (B/N)Ab ratios compared to adults (1.9 vs. 0.4 for ID_50_, P=0.0002). Findings suggest that young children mount robust antibody responses to SARS-CoV-2 following community infections. Additionally, these results support using neutAb to measure the immunogenicity of COVID-19 vaccines in children aged 0-4 years.

---

Our understanding of the epidemiology of SARS CoV-2 infection in children has evolved since late 2019. Early in the pandemic, SARS-CoV-2 infections were less frequently diagnosed in children than adults, raising questions about whether children were less susceptible to infection. Studies have since documented that children can be infected at similar rates as adults^1^ and can transmit infection.^2 3 4^ Although children can develop severe COVID-19, they are more likely than adults to be asymptomatic or mildly symptomatic,^5 6^ suggesting that the immunologic response to infection may vary with age. Data are mixed about whether children mount more robust SARS-CoV-2 antibody responses than adults following infection.^7 8 9^ Information about children aged 0-4 years is especially limited, with relatively small case series reported. While COVID-19 vaccines are recommended for children aged 5-17 years, evaluation of several vaccines in children aged 0-4 years is ongoing. Assessment of the magnitude and quality of the antibody responses to SARS-CoV-2 infection in very young children could inform COVID-19 vaccine assessment and deployment in this age group. Determination of the magnitude of SARS-CoV-2 receptor binding domain (RBD) antibody relative to neutralizing antibody may be useful, as a predominance of binding relative to neutralizing antibody has been observed in response to some other viral infections and vaccines.^10, 11 12^ Moreover, differences in binding to neutralizing antibody ratios have been observed when comparing responses to SARS-CoV-2 infection and vaccination.^13^

**S**ARS-CoV-2 **E**pidemiology **A**nd **R**esponse in **Ch**ildren (SEARCh) is a prospective household cohort study designed to address SARS-CoV-2 susceptibility, illness, transmission, and immunologic responses in children aged 0-4 years and their household members (Methods). In this cross-sectional analysis of enrollment sera, we compare titers of SARS-CoV-2 binding and neutralizing antibody to wild-type (wt)SARS-CoV-2 and the Delta variant in adults, children aged 5-17 years, and children aged 0-4 years.

Sera were collected from 682 SEARCh participants in 175 households, including 332 (49%) adults aged 18-62 years, 96 (14%) children aged 5-17 years, and 254 (37%) children aged 0-4 years. We detected RBD antibody (RBDAb) in sera from 55 (8%) participants in 22 households, including 27 RBDAb seropositive children (Supplementary Figure 1). The proportion of RBDAb seropositive participants that reported suspected COVID-19 prior to enrollment varied by age: 7/14 (50%) children aged 0-4 years, 5/13 (38%) children aged 5-17 years, and 21/28 (75%) adults (*P*=0.03 for adults vs. all children). In 8 households with >1 member with prior suspected SARS-CoV-2 infection, all suspected infections were reported to have occurred within one calendar week of each other. None of the participants was hospitalized with COVID-19 disease before enrollment.

Among the 55 participants who were RBDAb seropositive, the geometric mean titer (GMT) of RBDAb (binding antibody units [BAU]/mL) was 10-fold higher in children aged 0-4 years than adults (373 vs. 35, *P*<0.0001) (Figure 1a); children aged 5-17 years also had higher RBDAb GMT than adults (268 vs. 35, *P*=0.0001, Figure 1a). Results were similar when adults from households without SARS-CoV-2 seropositive children were excluded (373 vs. 42, *P*<0.0001; 268 vs. 42, *P*=0.002). Thirteen of 22 (59.1%) households had ≥1 member seropositive for RBDAb; all enrolled household members were seropositive in 4/22 households (18.2%) (Figure 1b). Children aged 0-4 years had the highest RBDAb titers in 11/12 households with a seropositive child aged 0-4 years (Figure 1b).

**Figure 1.**
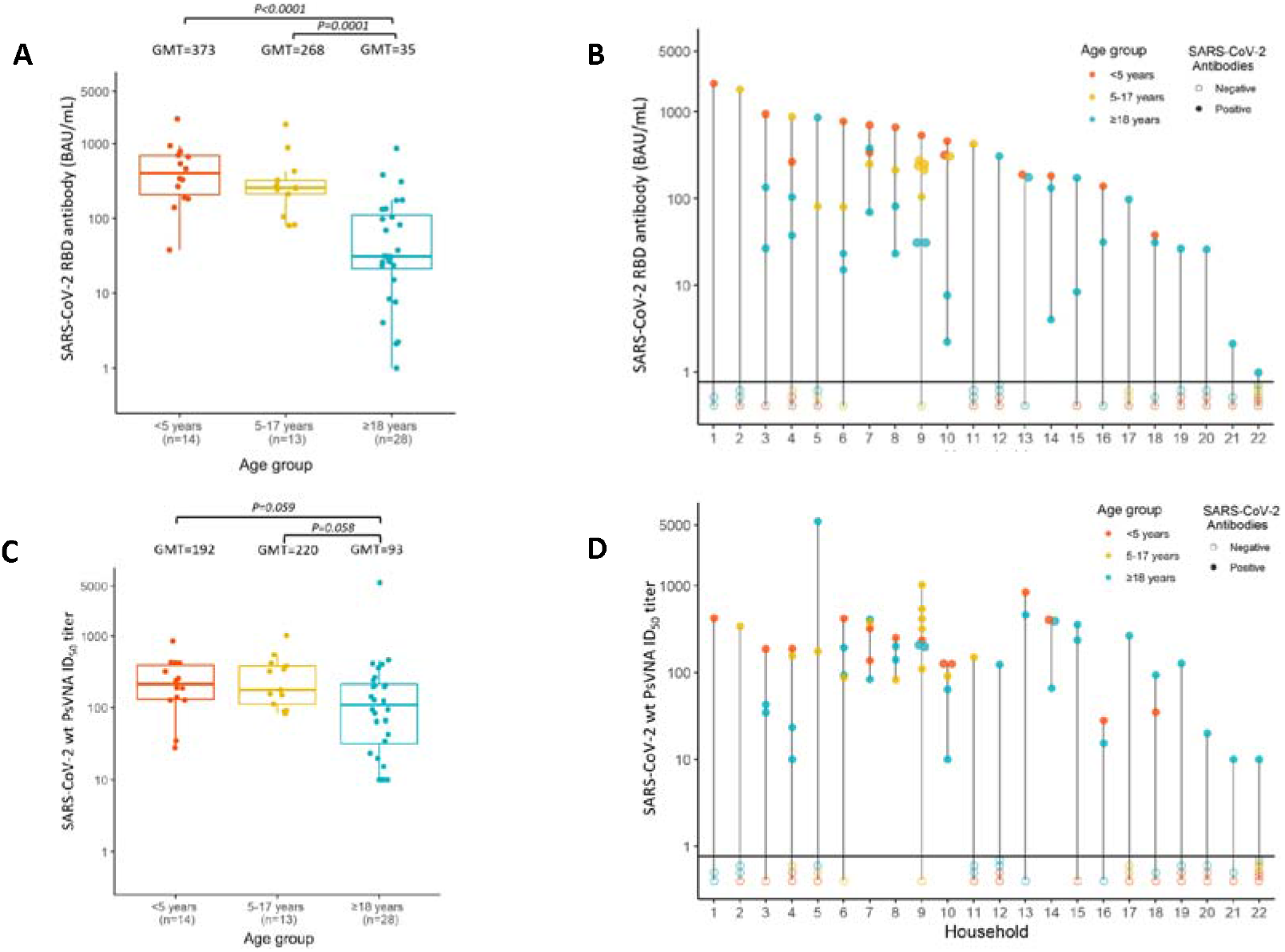
SARS-CoV-2 serum antibody titers at enrollment in SEARCh. **Figure 1a:** SARS-CoV-2 RBDAb titers by age, BAU/mL (orange= children 0-4 years, yellow= children 5-17 years, blue=adults). **Figure 1b:** RBDAb titers by household. Color scheme is the same as for 1A; filled circles represent RBDAb titers above the cutoff (seropositive) and open circles represent RBDAb titers below the cutoff (seronegative). **Figure 1c:** wt SARS-CoV-2 neutralizing antibody ID_50_ titers measured by pseudovirus viral neutralizing antibody (PsVNA); shown by age group as in Figure 1A. **Figure 1d:** PsVNA ID_50_ titers by household, using the sequence and color scheme shown for Figure 1B.

Children aged 0-4 and 5-17 years had similar pseudovirus viral neutralizing antibody (PsVNA) 50% inhibitory dilution (ID_50_) titers to wild type (wt) SARS-CoV-2 (Fig 1C). Overall, PsVNA ID_50_ titers among children aged 0-17 years were 2-fold higher than among adults (GMT 205 vs. 93, *P*=0.02) (Supplementary Figure 2). Children aged 0-4 years also had the highest PsVNA ID_50_ titers of all seropositive household members in 9/12 households (Fig 1d). PsVNA ID_80_ titers showed similar patterns by age and household (Supplementary Figure 3). PsVNA ID_50_ GMTs to the Delta variant of SARS-CoV-2 were comparatively modest in all age groups (children aged 0-4, 192 wt vs. 58 Delta; children aged 5-17, 220 wt vs. 71 Delta; adults, 93 wt vs. 34 Delta) and the ratio of titers to wt and the Delta variant did not differ between age groups (not shown).

The B/N GMT ratio (GMTR) using PsVNA ID_50_ was highest in children aged 0-4 years (1.9), followed by children aged 5-17 years (1.2), and lowest in adults (0.4; *P*=0.0002 for children aged 0-4 years vs. adults; *P*=0.008 for children 5-17 years vs. adults) (Figure 2a). Trends were similar for the B/N GMTR using PsVNA ID_80_ (Figure 2b).

**Figure 2.**
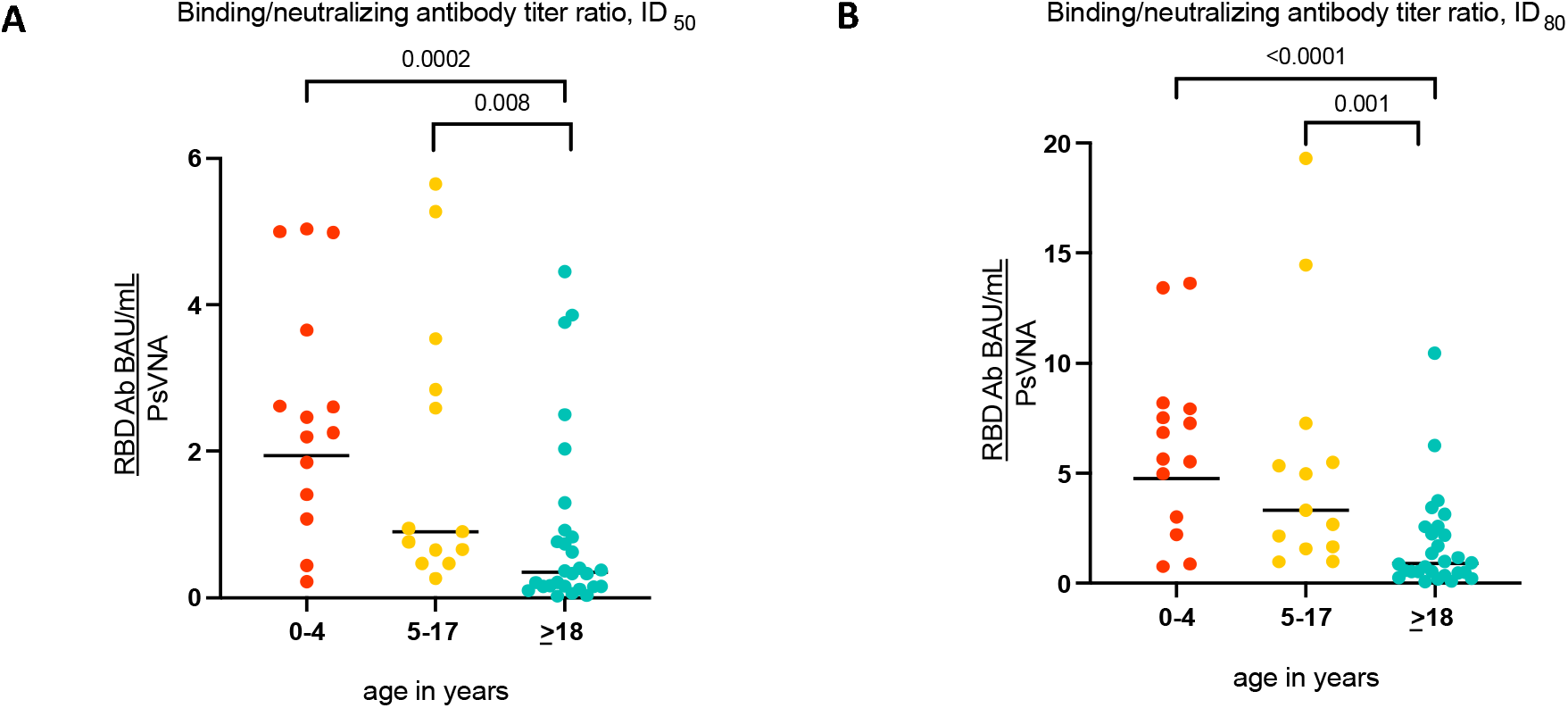
GMTR of SARS-CoV-2 RBDAb (BAU/mL) to SARS-CoV-2 PsVNA (orange= children 0-4 years, yellow= children 5-17 years, blue=adults 18-62 years). **Figure 2a:** ID_50_ titers. **Figure 2b:** ID_80_ titers.

In this analysis of 55 adults and children with serologic evidence of prior SARS-CoV-2 infection, children aged 0-4 years had ∼10-fold higher levels of RBDAb and ∼2-fold higher levels of neutralizing antibody to wt SARS-CoV-2 compared to adults. The consistency of these findings within households suggests that the differences were likely unrelated to timing of infection, as household members would likely have been infected with SARS-CoV-2 at approximately the same time. Differences in the relative magnitude of Ab response between children and adults were also unlikely to be from differences in illness severity ^9^, as many children had no known history of COVID-19, suggesting they experienced mild or subclinical infection.

Relatively few studies have directly compared the antibody responses to SARS-CoV-2 in children and adults, particularly between children aged 0-4 years and adults. A study of hospitalized patients found that adults mounted higher neutralizing antibody responses than children.^7^ In contrast, one community-based study of household clusters of mild COVID-19 found that children had higher and more sustained titers of SARS-CoV-2 neutralizing antibody than adults.^8^ Another community-based study found that children had higher titers of binding IgG antibody to SARS-CoV-2 spike, nucleocapsid, and RBD, and similar neutralizing antibody levels compared to adults.^9^ Our findings expand upon these community-based studies by 1) contributing additional data for children aged 0-4 years, a relatively understudied population with respect to immune responses to SARS-CoV-2; 2) demonstrating that differences in the magnitude of the RBDAb titer are consistent within households in which timing of infections are likely similar; and 3) demonstrating that although children generally develop higher titers of neutralizing antibody than adults, young children appear to make proportionally more SARS-CoV-2 RBDAb than neutralizing antibody, as evidenced by a higher B/N GMTR in very young children compared to adults. It is possible that these robust humoral immune responses diminish rates of serious or highly symptomatic infection by promoting viral clearance. Our findings are also consistent with the observation that children aged 5-11 years and adults develop comparable SARS-CoV-2 Ab responses to the BNT162b2 vaccine when children receive one-third of the dose given to adults.^14^

We observed a stepwise downward progression in B/N GMTR with age: children aged 0-4 years had the highest ratio of B/N Ab, children aged 5-17 years had an intermediate level, and adults had the lowest. The finding that young children had relatively high titers of binding antibody relative to neutralizing antibody was unexpected; the reasons for it are unknown. One possibility is that children aged 0-4 years have had fewer opportunities for priming infections with related betacoronaviruses^15^ and therefore may take longer to develop high-affinity neutralizing Ab than coronavirus-experienced older children and adults. The pattern of neutralizing Ab epitope recognition may also differ in young children and adults, as observed for other respiratory viruses.^11^ For example, adults may be more likely to recognize neutralizing epitopes in S2 or N-terminal domain than young children. Children may also have more durable RBD-specific Ab responses.^9^ The kinetics of the SARS-CoV-2 RBD and neutralizing Ab responses in each age group will be investigated using longitudinal samples obtained in the SEARCh study.

This study has several limitations. First, the number of SARS-CoV-2 RBDAb positive subjects was relatively small. Second, findings in this analysis likely largely reflect humoral immune responses to wt-like SARS-CoV-2 infections and may not be generalizable to infections with emerging variants. Third, this was a cross-sectional analysis and may not have fully accounted for potential differences in timing of infection between adults and children. However, data from households in which the timing of suspected COVID-19 infection was reported indicate that these events occurred in close temporal proximity. Additional longitudinal studies are needed to confirm our initial observations in larger groups of individuals in whom timing of infection is known and who were infected with other SARS-CoV-2 variants.

We have shown that young children aged 0-4 years who were infected with SARS-CoV-2 are capable of mounting substantial RBD binding and neutralizing Ab responses to wt SARS-CoV-2, and that their immune responses frequently exceeded those of adults in the same households. Our findings have two practical implications for COVID-19 vaccine development for the very young. First, they support the use of a reduced dose of the BNT162b2 vaccine in an ongoing trial in young children aged 0-4 years (Clinicaltrials.gov #04816643). Second, the differences in B/N GMTR by age suggest that RBDAb responses may not predict neutralizing Ab responses as reliably in young children as in adults,^16^ and that neutralizing Ab responses should continue to be assessed in vaccine trials involving this youngest age group.

## Data Availability

The data that support the findings of this study are not openly available due to reasons of sensitivity as they contain personal identifiable information, but are available from the corresponding author (RAK) upon reasonable request.

## Acknowledgements

We thank the Department of Pathology, Johns Hopkins University School of Medicine, for performing the SARS-CoV-2 Roche Elecsys assay, and the SEARCh families for participating in this study. This study was funded by the U.S. Centers for Disease Control and Prevention (CDC) Award 75D30120C08737.

## Methods

### Participants

Sera tested in this study were obtained at enrollment from individuals participating in SARS-CoV-2 Epidemiology And Response in Children (SEARCh), a longitudinal household-based cohort study in Maryland designed to examine the epidemiology and immune response to SARS CoV-2 infection. Households were eligible if they contained at least one child aged 0-4 years; individuals were eligible if they had not yet received a COVID-19 vaccine. SEARCh participants were enrolled between November 13, 2020 and February 27, 2021. Prior to study participation, all participants or their parents/guardians provided informed consent, and children aged 7 years and older also provided assent. Participants were then followed prospectively for evidence of SARS-CoV-2 infection using molecular and serologic diagnostic techniques for 8 months or through October 2021, whichever occurred sooner. These prospective data will be the subject of a future report. Participants or their parents/guardians were asked whether they were told by a healthcare provider that they had suspected or confirmed COVID-19 prior to enrollment. Household members also completed questionnaires throughout the study that included information on household composition, illness symptoms, and school, work, and leisure activities. Data were collected using Research Electronic Data Capture (REDCap). This study was funded by the U.S. Centers for Disease Control and Prevention (CDC) Award 75D30120C08737. The study protocol was reviewed and approved by the Johns Hopkins Bloomberg School of Public Health Institutional Review Board (IRB).

### Antibody assays

Enrollment sera were first tested for antibody to the receptor binding domain (RBD) of wild-type (wt) (WA1) SARS-CoV-2 using the Roche Elecsys Anti-SARS-CoV-2 S electrochemiluminescence immunoassay (Indianapolis, IN) in the Department of Pathology, Johns Hopkins School of Medicine. The correlation between the WHO standard and the Roche Elecsys Anti-SARS-CoV-2 is 0.9996 (Pearson’s r^2^) and the conversion factor between this assay and the WHO standard is 1.0288. RBD antibody titers are expressed as Binding Antibody Units (BAU)/mL. Sera that were RBD antibody positive were tested for neutralizing antibody using a pseudotyped reporter neutralization assay (PsVNA). Neutralization of wt (WA1.D614G) SARS-CoV-2 and of the Delta variant (B.1.617.2) of SARS-CoV-2 were measured at the Vaccine Research Center of the U.S. National Institutes of Health in a single-round-of-infection assay with lentivirus-based pseudotyped virus particles (pseudoviruses) as previously described. ^17 18^ The 50% and 80% inhibitory dilution (ID_50_ and ID_80_) titers are reported. ID50 is considered a more sensitive measure of neutralization, and is the customary metric in SARS-CoV-2 publications; ID80 is considered a more stringent measure. RBD antibody geometric mean titers (GMTs) and neutralizing antibody titers were compared by age group among all RBD Ab seropositive participants. Since SARS-CoV-2 infections may have occurred at any point prior to cohort enrollment and antibody titers may wane over time, individual RBD antibody and neutralizing antibody titers were also compared by age group within households, with the assumption that infections within households occurred at around the same time. The ratio of RBD binding antibody to wt neutralizing antibody, and the binding/neutralizing (B/N) GMT ratio (GMTR) were calculated for each participant and each age group.

### Statistical analysis

Analyses were performed using Graphpad Prism (San Diego, CA) and in R version .1.0 (R Foundation for Statistical Computing) in RStudio version 1.4.1717 (RStudio, Inc). Antibody titers were compared using the Mann-Whitney-Wilcoxon test (two-sided), and proportions were compared using Fisher’s exact test (two-sided). Figures were generated using Graphpad Prism and the Ggplot2 package.

### Code availability

Code used to generate figures will be made available upon request.

## Supplementary figures

**Supplementary Figure 1.**
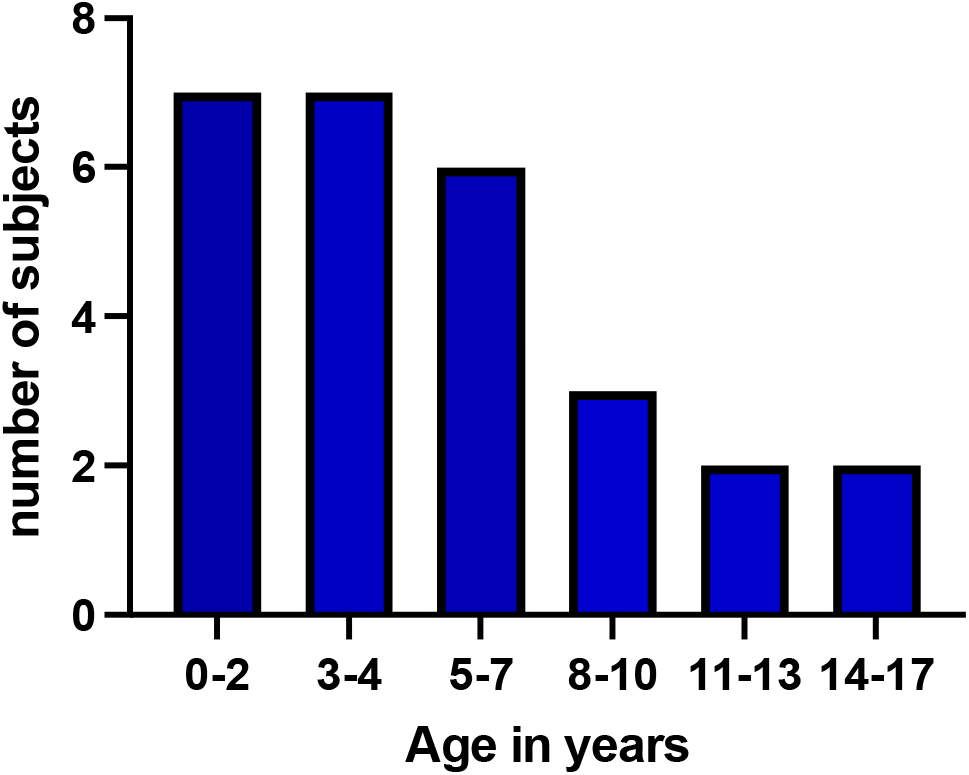
Age distribution of children in the SEARCh study who were SARS-CoV-2 seropositive at enrollment.

**Supplementary Figure 2.**
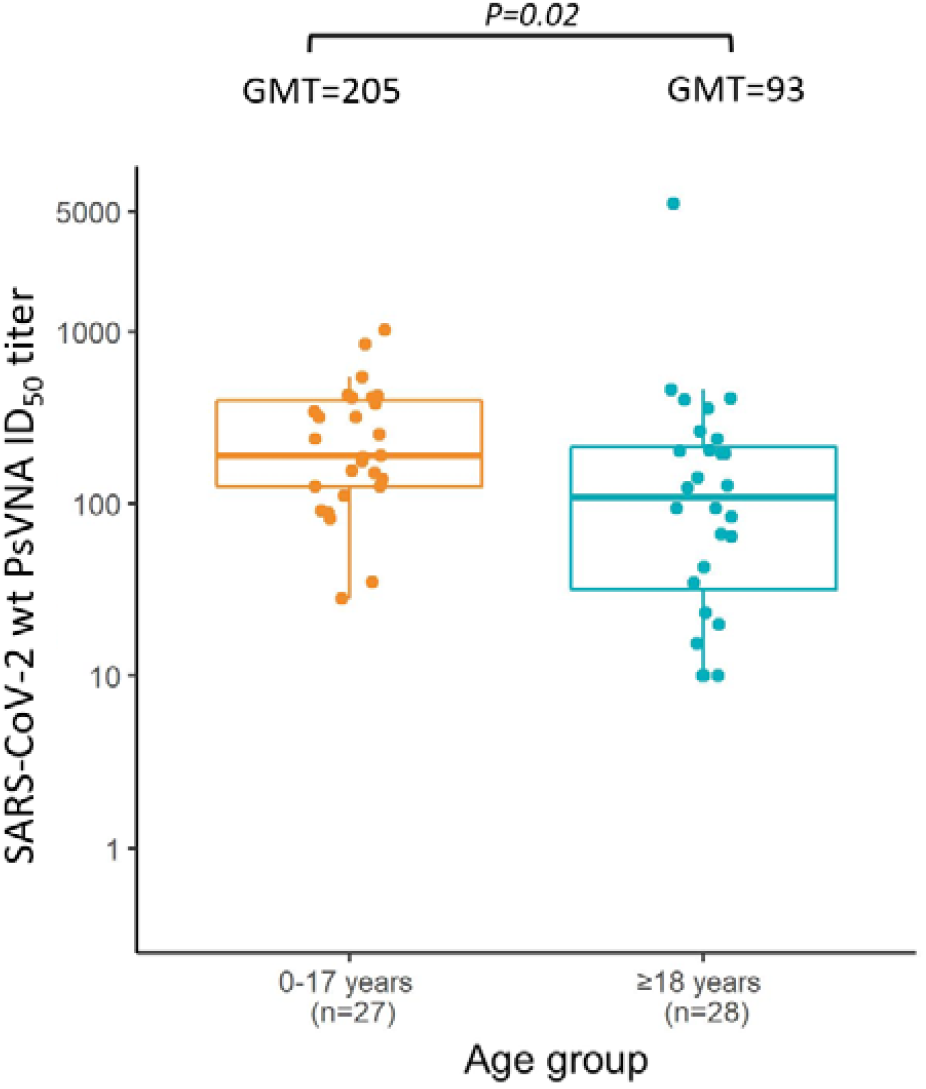
Box and whiskers plot of SARS-CoV-2 wt PsVNA ID_50_ titer for all children ages 0-17 (light orange) vs. adults (blue).

**Supplementary Figure 3.**
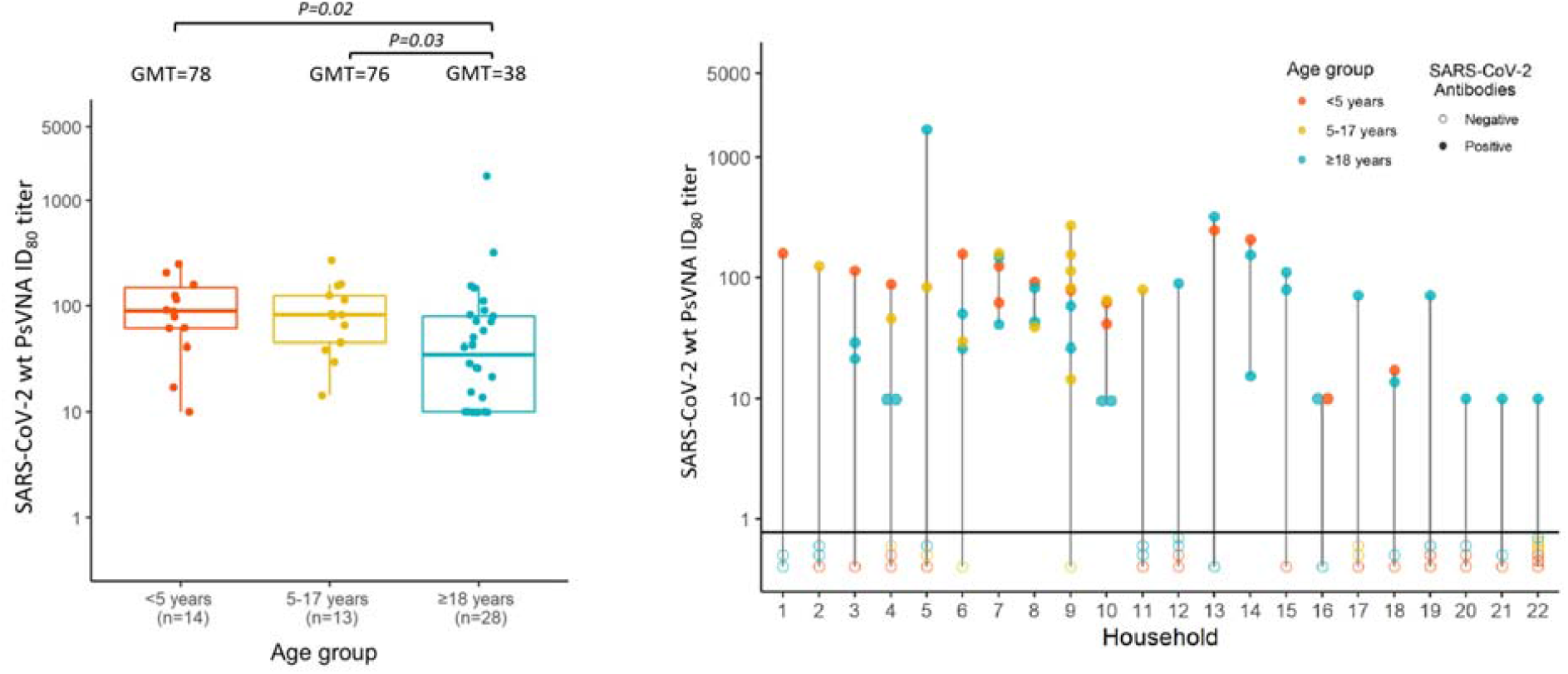
ID_80_ titers of neutralizing antibody to wt SARS-CoV-2, measured by PsVNA. **Fig.3a**, shown by age group, as in Figure 1a. **Figure 3b:** shown by household, as in Figure 1b.

## REFERENCES

1. Dawood, F.S., et al. Incidence Rates, Household Infection Risk, and Clinical Characteristics of SARS-CoV-2 Infection Among Children and Adults in Utah and New York City, New York. JAMA pediatrics (2021).

2. Paul, L.A., et al. Association of Age and Pediatric Household Transmission of SARS-CoV-2 Infection. JAMA pediatrics 175, 1151–1158 (2021).

3. Szablewski, C.M., et al. SARS-CoV-2 Transmission Dynamics in a Sleep-Away Camp. Pediatrics 147(2021).

4. Chu, V.T., et al. Household Transmission of SARS-CoV-2 from Children and Adolescents. The New England journal of medicine 385, 954–956 (2021).

5. Ladhani, S.N., et al. COVID-19 in children: analysis of the first pandemic peak in England. Archives of disease in childhood 105, 1180–1185 (2020).

6. Coronavirus Disease 2019 in Children - United States, February 12-April 2, 2020. MMWR Morb Mortal Wkly Rep 69, 422–426 (2020).

7. Pierce, C.A., et al. Immune responses to SARS-CoV-2 infection in hospitalized pediatric and adult patients. Science translational medicine 12(2020).

8. Bonfante, F., et al. Mild SARS-CoV-2 Infections and Neutralizing Antibody Titers. Pediatrics 148(2021).

9. Garrido, C., et al. Asymptomatic or mild symptomatic SARS-CoV-2 infection elicits durable neutralizing antibody responses in children and adolescents. JCI Insight 6(2021).

10. Hurtado-Monzón, A.M., et al. The role of anti-flavivirus humoral immune response in protection and pathogenesis. Rev Med Virol 30, e2100 (2020).

11. Goodwin, E., et al. Infants Infected with Respiratory Syncytial Virus Generate Potent Neutralizing Antibodies that Lack Somatic Hypermutation. Immunity 48, 339-349.e335 (2018).

12. Karron, R.A. Preventing respiratory syncytial virus (RSV) disease in children. Science (New York, N.Y.) 372, 686–687 (2021).

13. Amanat, F., et al. SARS-CoV-2 vaccination induces functionally diverse antibodies to NTD, RBD, and S2. Cell 184, 3936-3948.e3910 (2021).

14. Woodworth, K.R., et al. The Advisory Committee on Immunization Practices’ Interim Recommendation for Use of Pfizer-BioNTech COVID-19 Vaccine in Children Aged 5-11 Years - United States, November 2021. MMWR Morb Mortal Wkly Rep 70, 1579–1583 (2021).

15. Yang, H.S., et al. Association of Age With SARS-CoV-2 Antibody Response. JAMA Netw Open 4, e214302 (2021).

16. Salazar, E., et al. Convalescent plasma anti-SARS-CoV-2 spike protein ectodomain and receptor-binding domain IgG correlate with virus neutralization. The Journal of clinical investigation 130, 6728–6738 (2020).

17. Jackson, L.A., et al. An mRNA Vaccine against SARS-CoV-2 - Preliminary Report. The New England journal of medicine 383, 1920–1931 (2020).

18. Pegu, A., et al. Durability of mRNA-1273 vaccine-induced antibodies against SARS-CoV-2 variants. Science (New York, N.Y.) 373, 1372–1377 (2021).

